# Wastewater surveillance of influenza activity: Early detection, surveillance, and subtyping in city and neighbourhood communities

**DOI:** 10.1101/2022.06.28.22276884

**Authors:** Elisabeth Mercier, Patrick M. D’ Aoust, Ocean Thakali, Nada Hegazy, Jian-Jun Jia, Zhihao Zhang, Walaa Eid, Julio Plaza-Diaz, Pervez Kabir, Wanting Fang, Aaron Cowan, Sean E. Stephenson, Lakshmi Pisharody, Alex E. MacKenzie, Tyson E. Graber, Shen Wan, Robert Delatolla

**Affiliations:** Department of Civil Engineering, University of Ottawa, Ottawa, Canada, K1N 6N5; Children’s Hospital of Eastern Ontario Research Institute, Ottawa, Canada, K1H 8L1

## Abstract

Recurrent epidemics of influenza infection and its pandemic potential present a significant risk to global population health. To mitigate hospitalizations and death, local public health relies on clinical surveillance to locate and monitor influenza-like illnesses and/or influenza cases and outbreaks. At an international level, the global integration of clinical surveillance networks is the only reliable method to report influenza types and subtypes and warn of an emergent pandemic strain. During the COVID-19 pandemic, the demonstrated utility of wastewater surveillance (WWS) in complementing or even replacing clinical surveillance, the latter a resource-intensive enterprise, was predicated on the presence of stable viral fragments in wastewater. We show that influenza virus targets are stable in wastewaters and partitions to the solids fraction. We subsequently quantify, type, and subtype influenza virus in municipal wastewater and primary sludge throughout the course of a community outbreak. This research demonstrates the feasibility of applying influenza virus WWS to city and neighbourhood levels; showing a 17-day lead time in forecasting a citywide flu outbreak and providing population-level viral subtyping in near real-time using minimal resources and infrastructure.

## 1. Introduction

The World Health Organization (WHO) estimates that every year there are approximately 3,000,000 to 5,000,000 severe influenza infections causing between 290,000 and 650,000 deaths around the world^1^. In Canada, influenza is estimated to be the cause of approximately 12,200 hospitalizations and 3,500 deaths every year, making this infectious disease amongst the top 10 leading causes of death in the country^1,2^. The significant impacts of seasonal and pandemic influenza on global health, particularly in the context of the potential deregulation of the traditional seasonality of respiratory pathogens after the onset of COVID-19 has created an urgent need for an improved surveillance of this disease. Standard clinical surveillance is resource intensive and often lags behind community outbreaks. Furthermore, clinical surveillance also often provides data with insufficient region-specificity on flu activity and viral subtyping^3^. Wastewater surveillance (WWS) of influenza, due to the localized and high-enrollment nature of the test, has the potential to support and complement clinical surveillance programs and strengthen health emergency response systems in a similar manner to the demonstrated benefits of WWS for poliovirus observed during the 20^th^ century^4,5^ and currently with the application of wastewater surveillance of SARS-CoV-2, the virus responsible for COVID-19, during the COVID-19 pandemic^4,6–9^. WWS of influenza has the potential to be an anonymous, aggregated, economical, and rapid monitoring tool that captures a significant portion of communities through involuntary contributions and hence an important option for public health units/agencies that requires further development.

The influenza virus is a single-stranded RNA virus of the Orthomyxoviruses class and is divided into types A, B, C, and D. Influenza A virus (IAV) and influenza B virus (IBV) types are typically associated with seasonal influenza endemic activity, however different subtypes of IAV are also responsible for influenza pandemics^10,11^. Shedding characterization of IAV and IBV suggests the possibility for a viable application of WWS to identify and monitor seasonal influenza outbreaks and future global influenza pandemics^12^. Previous studies have reported elevated fecal shedding rates of IAV and IBV of up to 6 log^10^ copies/g^13^. In addition to high rates of shedding, infected individuals also maintain elevated fecal viral titers that are both higher and longer than that observed in detection periods of the virus in feces when compared to nasal swabs^14,15^.

Although IAV and IBV are fecally shed at a high rate, the virus is enveloped with an outer lipid membrane, and as a result, are often presumed to be present in low concentration in wastewater. Hence a thorough investigation of the partitioning of the influenza virus in wastewater and a subsequent optimized enrichment protocol of the influenza virus is required for the successful implementation of WWS. The hydrophobicity resulting from the outer lipid membrane suggests that IAV and IBV shall partition to the solids fraction of the wastewater matrix. In general, there is limited success in detecting endogenous IAV and IBV RNA in wastewater and largely partitioning experiments have been limited to the investigation of spiked concentrations of viral surrogates to wastewaters^16–18^. The fate and behavior of endogenous influenza virus and the targeted RNA that is likely present in low concentrations in wastewater is expected to differ from a spiked viral surrogate within a wastewater matrix. In this regard, preliminary investigations by Wolfe et al.^18^ recently hinted that RNA preferentially partitions to the solids fraction of wastewater compared to polyethylene glycol (PEG) precipitated supernatant, hence supporting the hypothesis of the disease target partitioning to the solids fraction of wastewater. As such, with the current knowledge of influenza partitioning in wastewater based predominantly on surrogate spiking experiments and on a single study based on endogenous IAV, additional partitioning studies performed on endogenous influenza virus in wastewater are necessary to optimize the enrichment of disease targets in the wastewater matrix and to apply influenza WWS within cities and neighbourhood communities.

Numerous studies of influenza virus in natural water sources have been undertaken to analyze the potential of oral influenza transmission pathways^17,19,20^. To date, only one study has outlined a protocol demonstrating the successful detection of influenza, IAV, in wastewaters^18^. This study recently demonstrated good agreement between wastewater measurements of IAV and reported clinical IAV cases observed as part of the Michigan University’ s campus surveillance program and Stanford University’ s athlete surveillance programs. However, a lack of information in regards to the quantification and trending of IAV WWS signal in cities and neighbourhood communities and most importantly the relation of the IAV signal in wastewater to clinical surveillance metrics. New information related to IAV WWS is hence required to elucidate complementary aspects of influenza virus WWS relative to clinical surveillance.

Furthermore, no studies to date demonstrate an ability for WWS to provide IAV subtyping information. Subtyping of IAV in wastewater is potentially especially useful to public health units/agencies to identify the presence/absence of IAV strains responsible for community disease during seasonal epidemic activity. Because IAV may undergo antigen shift or drift which favors pandemic-potential influenza features, it is of first importance to develop protocols and methodologies to subtype the virus in wastewater. Even with 18 hemagglutinin (H1 to H18) and 11 neuraminidase (N1 to N11) subtypes being known to exist for IAV, the subtypes that routinely cause human infections today are restricted to two strains, H1N1 and H3N2^11^. As such, identification of which of these two subtypes are responsible for community disease, with each subtype being associated with distinct clinically relevant phenotypic characteristics, will further increase the impact of IAV WWS programs.

Upon this background, and with the goal of advancing influenza WWS, we first investigated the partitioning behavior of endogenous IAV in city and neighbourhood wastewater. The partitioning findings were used to optimize an enrichment protocol for the quantification of influenza RNA in wastewater and subsequently apply the protocol to quantitate wastewater IAV and IBV RNA at the citywide level as well as within three distinct neighbourhoods in Ottawa, Ontario, Canada. The wastewater viral signal was correlated with clinical case data available from the Ottawa public health unit/agency during an out-of-season outbreak, determining the benefits and complementary attributes of influenza virus WWS at the citywide and neighbourhood level. Finally, IAV positive citywide and neighbourhood wastewater were tested for H1N1 and H3N2 to identify the circulating subtype responsible for community disease. To the best of our knowledge, this study is the first to demonstrate the applicability of influenza virus WWS and subtyping at the citywide and neighbourhood-level during an influenza community outbreak and the relation of influenza virus WWS and influenza wastewater subtyping to clinical surveillance.

## 2. Methods

### 2.1. Site descriptions

The wastewater samples collected in this study were harvested from the City of Ottawa’ s Robert O. Pickard Environmental Centre, its sole water resource recovery facility (WRRF), which processes wastewater from approximately 910,000 individuals, or approximately 91% of the city’ s population. In addition, wastewater samples were harvested from three manholes that enabled access to the city sewer collectors. These three sewer locations geographically isolate three distinct city neighbourhoods that were previously identified as vulnerable communities for COVID-19 due to the higher concentration of residents living and a higher percentage of residents working in congregate care settings (e.g., long-term care facilities). Further information about the sampling locations is provided in Table S1.

### 2.2. Wastewater sample collection, enrichment, and nucleic acid extraction

To obtain samples representative of the citywide viral load of IAV and IBV, hourly, 24-hour composite samples of primary clarified sludge were collected daily from the Ottawa WRRF between February 2, 2022, and May 24, 2022. To obtain samples representative of neighbourhood-level viral load of IAV and IBV, hourly, 24-hour composite municipal sewer wastewater samples were harvested two to three times a week from the studied neighbourhoods across the same period. Upon collection, the primary sludge samples were immediately refrigerated at 4°C at the WRRF to await transport and were placed in coolers with ice packs for transport to the laboratory for analysis. Harvested neighbourhood samples were immediately placed in coolers with ice packs and transported to the laboratory where they were then allowed to settle for 60 minutes, followed by decanting to obtain settled solids. 40 mL of well-homogenized primary sludge (WRRF samples) or 40 mL of the settled solids (neighbourhood sewer samples) was centrifuged and 250 mg of pelleted material was then extracted using the RNeasy PowerMicrobiome (Qiagen) methodology as previously described by D’ Aoust et al.^21^.

### 2.3. Wastewater sample RT-qPCR analysis

Measurements of IAV, IBV, pepper mild mottle virus (PMMoV), and SARS-CoV-2 were performed via singleplex RT-qPCR (Bio-Rad, Hercules, CA) using previously developed and described assays^22–25^. All primers and probes, PCR cycling conditions, and reagent concentrations are described in detail in Table S3. All samples were run in technical triplicates with non-template controls and 5-point standard curves prepared with an RT-ddPCR-quantified lab-propagated Hong Kong/1/68/MA/E2 (H3N2) strain of IAV (IAV positive control), and with the EDX RPPOS standard (Exact Diagnostics) (IBV positive control). The assay’ s limit of detection (ALOD) and quantification (ALOQ) for IAV’ s M gene region were approximately 3.5 copies/reaction and 5.7 copies/reaction, respectively. PCR efficiency ranged from 91–102% and R^2^ values were greater than 0.98 (n = 10) throughout the study.

### 2.4. Primary sludge and municipal wastewater fractionation experiments

Primary sludge collected from the city of Ottawa and municipal wastewater collected from three Ottawa neighbourhoods were used to determine the fractionation of endogenous influenza RNA in these two wastewater matrices. As this study was performed during an outbreak of IAV activity in the city of Ottawa during the spring of 2022, the fractionation experiments were performed on IAV positive primary sludge and municipal wastewater samples.). All replicates were subject to the same storage, transport, and holding times prior to the fractioning experiments. All fractions were extracted within 2 hours of the fractionation procedure.

Two 250 mL primary sludge samples were collected on different days and subsequently split into 3 biological replicates (n=6) and then separated into three fractions: settled solids, PEG-precipitated solids, and supernatant (Fig. 1 A). To study primary sludge fractioning, samples were completely mixed, and 30 mL of sludge was collected and centrifuged at 10,000 x g for 45 minutes at 4°C. The supernatant was decanted and set aside, being careful to preserve the pellet. Samples were again centrifuged (10,000xg, 10 minutes, 4°C) and the remaining supernatant was then decanted and set aside. The remaining pellet was then stored at 4°C until extraction. The pellet sample fraction was considered the settled solid fraction of the sample and was extracted using the RNeasy PowerMicrobiome Kit (Qiagen), as previously described by D’ Aoust *et al*.^8^. The retained supernatant (∼27 mL) was serially filtered through 30 kDa-15 mL Amicon ultrafiltration cartridge (EMD Millipore) at 4,000 x g for 15 minutes at 4°C. This supernatant fraction was then extracted using the QIAamp Viral RNA Mini Kit (Qiagen) on a QIAcube Connect automated extraction platform as per the manufacturer’ s instructions. Finally, 30 mL of sludge was precipitated with a PEG 8000 solution at a final concentration of 80 g/L, 0.3M NaCl, and adjusted to a pH of 7.3, in a final volume of 40 mL. The samples were then mixed and incubated overnight at 4°C. The sample was then centrifuged at 10,000 x g for 45 minutes at 4°C. Samples were again centrifuged at 10,000 x g for 10 minutes at 4°C. The remaining pellet was then stored at 4°C until extraction. This sample fraction was considered the PEG-precipitated solids fraction of the sample and was extracted using the RNeasy PowerMicrobiome Kit (Qiagen), following previously described methodologies by D’ Aoust *et al*.^21^.

**Fig. 1.**
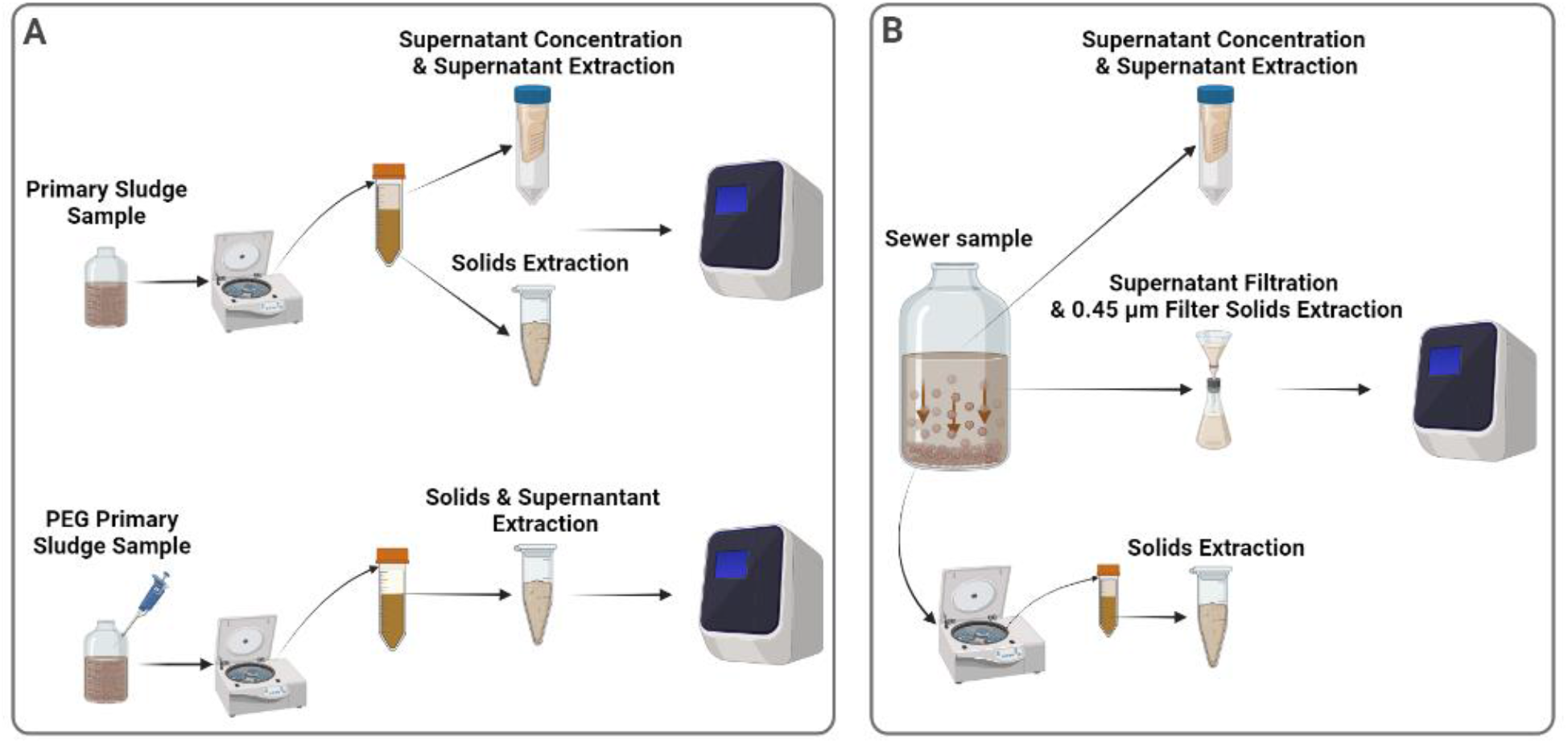
Schematic showing sample processing for the fractionation experiments of both types of tested wastewater. **A** Processing of primary clarified sludge samples to examine the fractionation of influenza A viral signal within the supernatant and solid pellet with and without PEG addition. **B** Processing of municipal wastewater to examine the fractionation of influenza A viral signal within the supernatant, filtered suspended solids and solid pellet.

To determine the fractionation of endogenous IAV in municipal wastewater, three 4.0 L municipal wastewater samples were collected on different days from the three neighbourhoods in Ottawa, split into 4 biological replicates (n=12), and subsequently separated into three fractions: settled solids, 0.45 µm filter solids, and supernatant (Fig. 1 B). For the municipal wastewater fractioning experiment, samples were settled for 2 hours at 4°C and then 40 mL of the settled solids were harvested and centrifuged at 10,000 x g for 45 minutes at 4°C. The supernatant was decanted and set aside, being careful to preserve the pellet. Samples were recentrifuged at 10,000 x g for 10 minutes at 4°C and the remaining supernatant was again set aside. The final pellet, which was considered the settled solid fraction, was stored at 4°C until extraction, using the RNeasy PowerMicrobiome Kit (Qiagen), as previously described by D’ Aoust *et al*.^8^. Subsequently, 40 mL of the post-settling supernatant of each sample was serially filtered through the 30 kDa-15 mL Amicon cartridge at 4,000 x g for 15 minutes at 4°C to generate the supernatant fraction of the sample. This was extracted using the QIAamp Viral RNA Mini Kit (Qiagen) on a QIAcube Connect automated extraction platform as per the manufacturer’ s instructions. Finally, 500 mL of supernatant (post-settling) of each sample was serially filtered through a 1.5 µm glass fiber filter (GFF) followed by filtration through a 0.45 µm GF6 mixed cellulose ester (MCE) filter (EMD Millipore). 32 mL of elution buffer (0.05 M KH2PO4, 1.0 M NaCl, 0.1% (v/v) TritonX-100, pH 9.2) was then passed through the spent filters (1.5 µm and 0.45 µm). The resulting eluate, considered the 0.45 µm filter solids fraction of the sample, was captured and stored at 4°C until extraction using the RNeasy PowerMicrobiome Kit (Qiagen), as previously described methodologies by D’ Aoust *et al*.^8^.

It should be noted that several enrichment methods were used in this study to partition the primary sludge and municipal wastewaters. In particular, the study applies PEG precipitation, filtration, and centrifugation, as these methods have been shown to appropriately concentrate enveloped viruses in wastewater^26–36^. In addition, it is noted that two RNA extraction kits were used to analyze the distinct solids fractions and the distinct liquid fractions of the primary sludge and municipal wastewater samples. The RNeasy PowerMicrobiome Kit (Qiagen) extraction kit was used to extract solids-rich fractions and has been previously demonstrated for the preferential extraction of viral RNA from solids. The QIAamp Viral RNA Mini Kit (Qiagen) extraction kit was used to extract liquid fractions and has been demonstrated for the preferential extraction of liquid fractions of wastewaters^37–40^.

The percentage of IVA gene copies partitioned to the primary sludge solids fraction was calculated by multiplying the gene copies measured in the extracted mass of solids by the ratio of the total mass of pelleted solids to the extracted mass. PEG precipitation was applied to the primary solids to calculate the viral signal present in the unsettled/suspended solids of the liquid fraction that were not detected by analyzing the liquid fraction alone. Thus, the IAV viral signal in the PEG-precipitated solids fraction was calculated by subtracting the IVA gene copies measured in the solids precipitated via the application of PEG from the IVA gene copies measured in the primary sludge solids fraction without the application of PEG. Finally, the IVA gene copies in the primary sludge supernatant fraction were calculated by simply multiplying the gene copies measured in the extracted liquid phase by the ratio of the total liquid volume to the extracted volume. The total measurable endogenous IAV signal from the primary sludge samples was subsequently calculated by summing the IVA gene copy fractions. The partitioning of the total signal was expressed as the percentage of the total measurable signal of the fractions.

The amount of IVA gene copies in the municipal wastewater solids fraction was calculated by multiplying the gene copies measured in the extracted mass of solids by the ratio of the total mass of pelleted solids to the extracted mass. The IVA gene copies in the 0.45 µm filtered solids fraction and supernatant fraction was calculated by multiplying the gene copies measured in the extracted sample by the ratio of the total sample volume to extracted volume. The total measurable endogenous IAV signal from the municipal wastewater samples was calculated by adding the three IVA gene copy fractions together. The partitioning of the total signal was again expressed as a percentage of the total measurable signal.

### 2.5. Clinical data

All influenza clinical data was provided weekly by Ottawa Public Health. Influenza screening of patient clinical samples in Ottawa is performed via RT-PCR assays either at the regional or provincial level, depending on whether patient samples originate from hospital inpatient testing or institutional outbreaks, respectively. Testing criteria for influenza screening in Ontario include: i) pediatric (<18 years old) emergency room patients with respiratory infection symptoms, ii) all hospitalized inpatients showing signs and symptoms of respiratory infection, iii) patients showing signs and symptoms of respiratory infection in institutions where there has been a declared outbreak, and iv) patients showing signs and symptoms of respiratory infection in institutions where there has not been a declared outbreak^41^. Clinical samples of type 3 and 4, namely the institution patients’ samples, are the only ones sent to Public Health Ontario Laboratories (PHOL) where they are typed and subtyped. All other samples are sent to the Eastern Ontario Regional Laboratory Association (EROLA) to be typed only.

### 2.6. Statistical analyses

Time-step Pearson’ s R correlation analyses were performed using Graphpad Prism (version 9.3.1) to evaluate the fit between observed IAV viral signal in wastewater and reported IAV clinical data obtained from the local public health unit (Ottawa Public Health) and to determine the lag/prediction period between observed IAV viral signal in wastewater and reported IAV clinical cases at various times (0 to 21 days time-stepped). Normality of the data was established beforehand with a quantile-quantile plot to determine the validity of using Pearson’ s R correlation.

## 3. Results

### 3.1. Partitioning of endogenous IAV virus in primary sludge and municipal wastewaters

The city of Ottawa and its neighbourhoods experienced an uncommon seasonal outbreak of IAV activity in the spring of 2022. This IAV activity in the community also corresponded with a SARS-CoV-2 resurgence, with the two waves being likely initiated by the removal of the masking mandates in the city during the COVID-19 pandemic. IBV was not detected in clinical or wastewater samples in the city or the neighbourhoods during the resurgence of the viral diseases in the community. As such, endogenous IAV fractionation experiments were performed on primary sludge (citywide samples) and municipal wastewater (neighbourhood samples) as endogenous IBV primary sludge and municipal wastewater samples were unavailable.

In primary sludge, IAV was found to almost exclusively concentrate in the solids fraction; analysis of primary sludge detected 88.1 ±10.6% of the viral signal in the settleable solids with <0.1% present in both PEG-precipitated solids and liquid fractions (Fig. 2A). Similar partitioning was observed in municipal wastewater; 84.6 ± 10.3% of the viral signal was detected in the settleable solids, 4.0 ± 3.1% was present in suspended solids larger than 0.45 µm (filtered), and <0.1% found in the supernatant (Fig. 2B). These findings are in agreement with the only other modern study, similarly reporting an IAV tropism for settled solids in wastewater^19^. Based on the findings of the fractionation experiments performed for IAV, primary sludge, and municipal wastewater sample processing and viral signal measurements for IAV in this study were exclusively performed using solids-optimized enrichment and extraction methodologies.

**Fig. 2.**
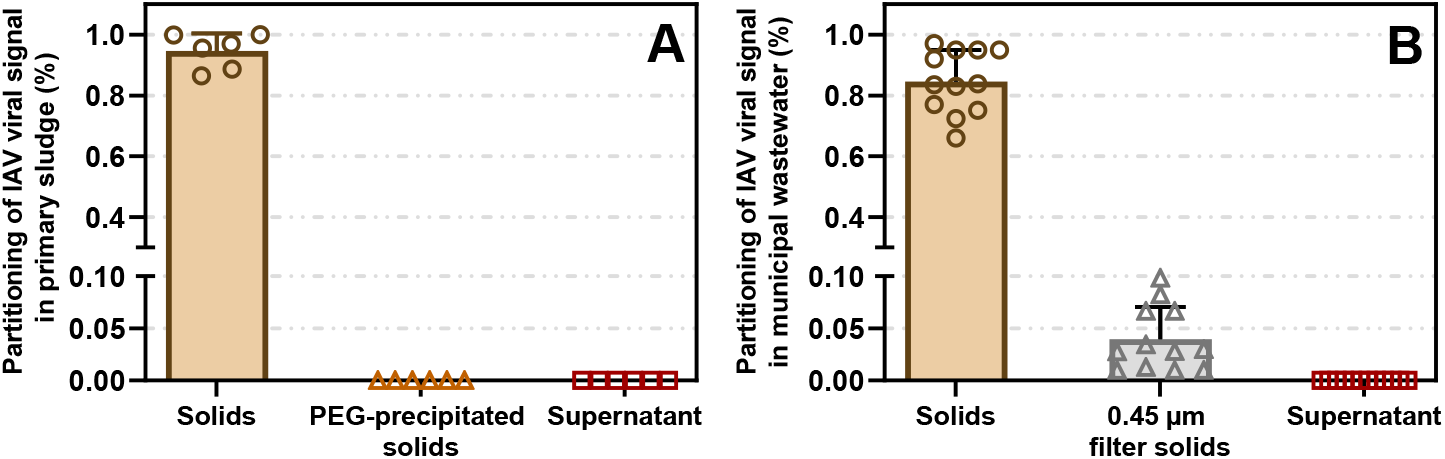
Results of fractionation experiments of IAV viral signal in wastewater. Partitioning of the measured endogenous IAV viral signal present in: **A** primary sludge (n=6); **B** municipal wastewater (n=12). Mean and standard deviation are displayed. Where the standard deviation is too small, the error bars are not displayed. Each measurement is done in 3 technical triplicates form each biological replicates.

### 3.2. Surveillance of wastewater IAV & IBV RNA at the citywide level and in neighbourhoods

IAV was detected in 79 (60%) of the 131 samples tested, while IBV was detected in none. Sanger sequencing of PCR products confirmed the presence of IAV in primary sludge in samples from both WRRF and the three neighbourhoods sampled. Aligned sequences of the PCR products from primary sludge and municipal wastewater samples showed 92.5% and 98.1% homology with IAV (H3N2, Hong Kong/1/68/MA/E2 subtype), respectively. Meanwhile, SARS-CoV-2 viral signal above the limit of quantification^21^ was detected in 123 out of the 131 samples tested (94%). IAV and SARS-CoV-2 signals were normalized using PMMoV signal to account for the human fecal content of the wastewater solids (Fig. 3). PMMoV has been recommended as a suitable normalizing factor enabling identification of trends in part due to stability in wastewater and low temporal variation in concentration, particularly when analyzing the solid fraction of wastewater^8,24,42–47^.

**Fig. 3.**
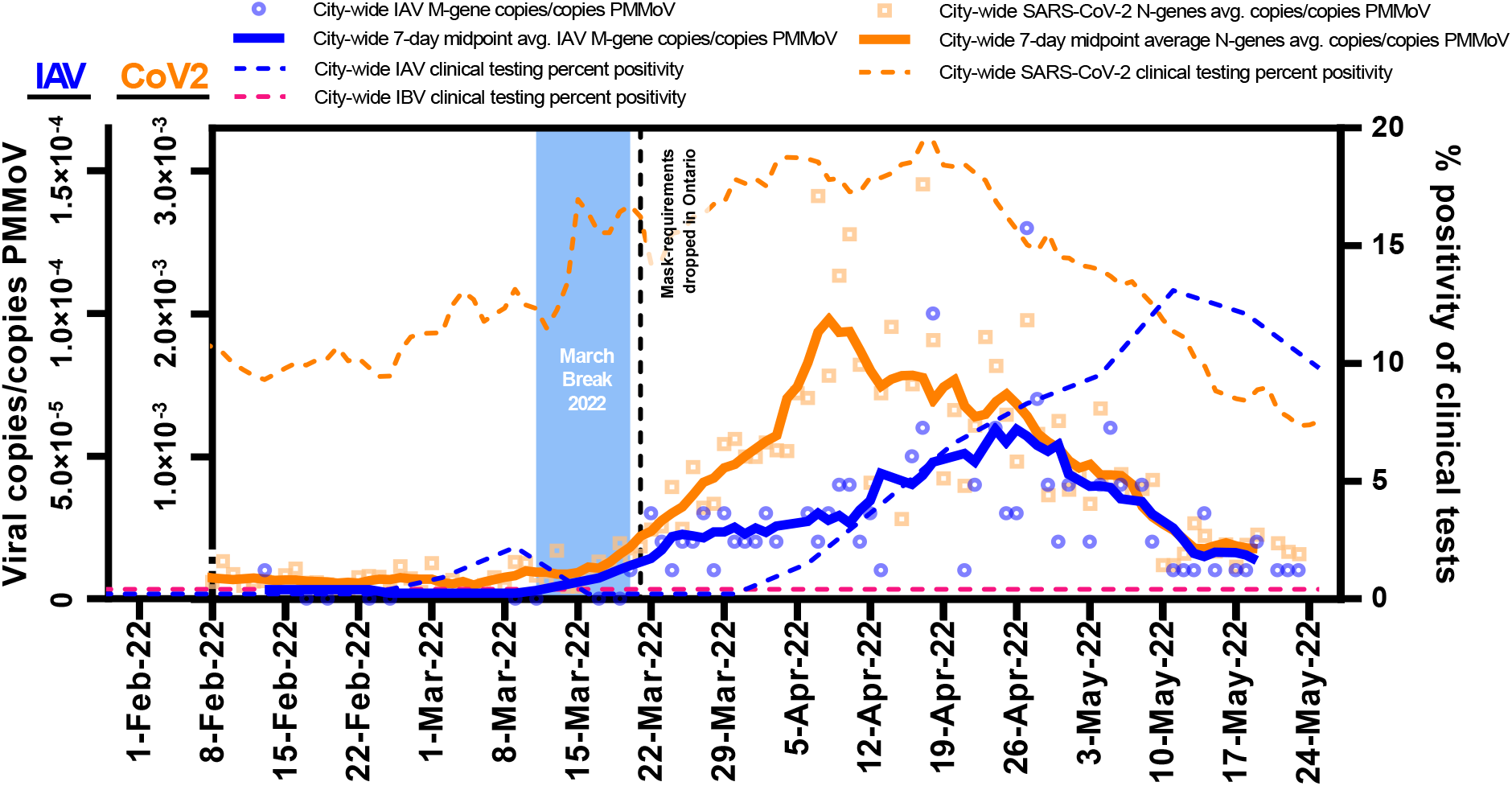
Comparison of1 IAV and SARS-CoV-2 wastewater signals over time at the citywide level. A comparison of both IAV and SARS-CoV-2 wastewater signals at the citywide level in Ottawa, from samples harvested from the city’ s WRRF. It was demonstrated that the detection of IAV in wastewater at the city-level (Feb. 13, 2022) occurred 17 days before the first clinical detection of IAV at any of the city’ s hospitals or clinics.

Mandated masking requirements in the province of Ontario, which were applied during the COVID-19 pandemic, were largely lifted on March 21, 2022. The lifting of the mandate combined with a one-week student March break (ending March 21, 2022) coincided with a predictable increase in the 7-day average PMMoV-normalized concentration of SARS-CoV-2 along with the initiation of an IAV wave in the city (Fig. 3). The citywide IAV signal did not increase as markedly as SARS-CoV-2 during the same post-masking protection mandate resurgence. This distinction between the rates of increase of IAV and SARS-CoV-2 signal in the city can likely be attributed to various factors such as population susceptibility to the two diseases, transmissibilities of the diseases, incubation periods of the diseases, and possible differences in degradation rates of the disease targets within the sewer system and within primary sludge clarifiers. IAV typically has a shorter mean incubation period (2.0 days)^48–50^ compared to ancestral SARS-CoV-2 (6.4 days)^51,52^, parainfluenza (4.0 days)^50^, respiratory syncytial virus (5.0 days)^50^, SARS-CoV-2 Delta variant (4.8 days)^53,54^ and Omicron variant (3.6 days)^53,54^. Interestingly, the decline of both wastewater viral signals was observed at a similar rate and ended on similar dates. The similarity in the trend may simply be due to the end of the flu season coinciding with the exhaustion of Omicron BA.1 in the city of Ottawa. Alternatively, population-wide behavior may have imparted a similar impact on the transmission of both viruses; regardless of which postulation is correct, the observed overlap of the virus wastewater profiles argues against the phenomenon of viral interference, the presence of one virus diminishing the spread of the other^55^.

Citywide PMMoV-normalized IAV concentrations in primary sludge showed moderate to strong correlation with weekly IAV clinical positivity rates (Pearson’ s r = 0.50, p <0.05, n = 14). It should be noted that this moderate to strong correlation was observed without the application of a time-step shift applied to the IAV wastewater signal, which visually precedes the clinical cases. A time-step correlation analysis revealed that when shifting the IAV wastewater signal 17 days forward, the Person’ s R correlation between the wastewater signal and clinical data indeed increased to a remarkably strong correlation (r = 0.97, p <0.05, n = 14). The identified overall lead time of the IAV wastewater signal of 17 days was identified as the highest Person’ s R correlation compared to time-steps tested between 0 to 21 days. The specific lead times of WWS over clinical testing related to the first detection in the wastewater, outbreak detection, peak, and resolution, ranged from 15–21 days (Table 1).

**Table 1.** Lead time of normalized IAV viral RNA signal weekly IAV percent positivity

| Outbreak progression | Lead time of wastewater to clinical metric | Details |
| --- | --- | --- |
| **First detection** | 17 days | WWS: First identification of IAV in population.<br>Clinical: First identification of IAV in population. |
| **Outbreak detection** | 21 days | WWS: First 3 consecutive days of increasing IAV viral signal<br>Clinical: First 2 consecutive weeks of increasing weekly percent positivity |
| **Peak of outbreak** | 15 days | WWS: Date of highest 7-day midpoint avg. IAV viral signal<br>Clinical: Date of highest percent positivity |
| **Outbreak resolution** | 17 days | WWS: 7 consecutive days of decreasing 7-day midpoint avg. IAV viral signal<br>Clinical: 2 consecutive weeks of decreasing weekly percent positivity |

Several factors can explain the extensive lead time of WWS. First, IAV clinical testing in Ottawa, is regulated by Public Health Ontario and typically prescribed when patients meet one of the four conditions described in the methods are met: i) pediatric (<18 years old) emergency room patients with respiratory infection symptoms, ii) all hospitalized inpatients showing signs and symptoms of respiratory infection, iii) patients showing signs and symptoms of respiratory infection in institutions where there has been a declared outbreak, and iv) patients showing signs and symptoms of respiratory infection in institutions where there has not been a declared outbreak^41^. Furthermore, the clinical testing data is reported at the citywide level by the date of clinical testing and not by the onset of symptoms, thus making it likely that fecal shedding of viral RNA could begin before individuals begin seeking medical treatment and being clinically tested. Additionally, due to the well-documented spread of many diseases in younger children in early-childcare centers and schools^56–59^, IAV can likely infect large numbers of individuals (young children and their parents) relatively undetected by current clinical surveillance, before transitioning to more vulnerable population groups (e.g., the elderly, sick or immunocompromised individuals) who may find themselves in greater need of medical treatment, and more likely to undergo clinical testing. Overall, this demonstrates that WWS more accurately reflects the onset of illness in the studied community compared to clinical surveillance and hence WWS can serve as an invaluable adjunct to clinical surveillance, informing the implementation of appropriate public health and patient management strategies.

Three neighbourhoods were also monitored for both IAV and SARS-CoV-2 viral signals in wastewater (Fig. 4). The neighbourhoods and their sewershed characteristics are described in greater detail in Table S1. At the neighbourhood level, peaks of both IAV and SARS-CoV-2 viral signals were observed earliest in neighbourhood #1 compared to the other neighbourhoods and compared to the citywide signal. The mean age of residents was significantly lower in this community (approximately 10 years younger) as compared to the other two neighbourhoods or compared to the city of Ottawa at large. It is hypothesized that the lower mean age of this neighbourhood could indicate the presence of more children, which could have contributed to an earlier transmission of IAV, allowing IAV viral signal in wastewater to peak in this neighbourhood before others or before the city’ s WRRF, particularly in the context of lower vaccination rates for young children^60^. This neighbourhood saw significantly more elevated viral signal levels for both IAV and SARS-CoV-2 on several occasions, supporting the idea that more in-community transmission of both viruses was occurring during the studied period. The IAV viral signal in neighbourhood #2 did not appear to deviate significantly from the citywide signal observed in primary sludge. Coincidentally, this neighbourhood Is characterized by a population age and number of households closest to the citywide average, perhaps hinting at a similar population distribution. The onset of the IAV outbreak in this neighbourhood was first detected on March 16, 2022, which was after the first detection of clinical cases of IAV at the city-wide level. The most elevated PMMoV-normalized IAV viral signal was observed in neighbourhood #3. It is hypothesized that because neighbourhood #3 has both the highest population and the fewest number of households of all studied neighbourhoods (Table S1), more congregate living situations may exist, which could have led to more community transmission of IAV, a trend often observed in locations experiencing higher population density^61–63^. Overall, these findings illustrate that detecting both IAV and potentially IBV in sewage samples collected from local, smaller geo-spatial areas may be used as an effective and economical strategy to track the timing, location, and magnitude of influenza activity outbreaks.

**Fig. 4.**
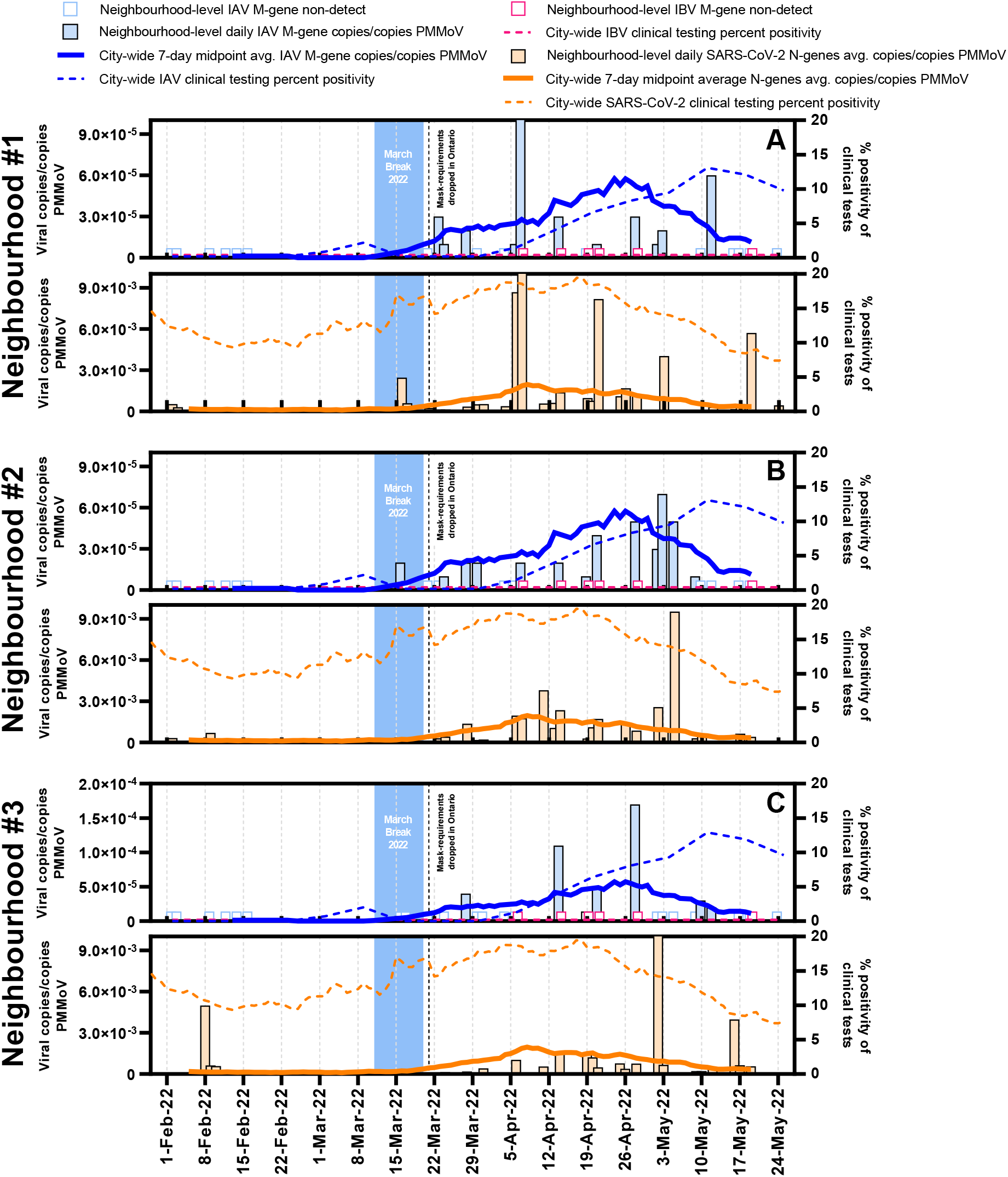
Comparison of2 IAV and SARS-CoV-2 wastewater signals over time at the neighbourhood level. A comparison of both IAV and SARS-CoV-2 wastewater signals at the neighbourhood level in Ottawa, from samples harvested from the sewer system. It was demonstrated that the normalized IAV viral signal differed between neighbourhood and with the citywide signal when population characteristic differed.

### 3.3. Subtyping of wastewater IAV RNA at the citywide level and in neighbourhoods

During the spring of 2022, the city of Ottawa experienced its first seasonal outbreak of influenza since the beginning of the COVID-19 pandemic. Through wastewater surveillance and clinical testing, it was determined that this outbreak was caused exclusively by IAV, and no IBV viral signal was detected in the wastewater or clinical samples. As such, the wastewater samples were retrospectively subtyped for IAV and hence screened for H1N1 and H3N2 at a rate of 1 sample per week. H3N2 was found to be the circulating IAV subtype in both the neighbourhoods and city-level samples, with no detection of H1N1. H3N2 was identified at the citywide level in the primary sludge sample via RT-qPCR on February 13, 2022, 17 days earlier than the appearance of the first clinical IAV case or the subsequent clinical subtyping of the virus. In Eastern Ontario, only two out of the four eligible sample categories, samples collected from the first four patients of an institution with a respiratory illness outbreak and samples from patients in institutions, not in an outbreak, are sent to Public Health Ontario Laboratories (PHOL) where they are typed and subtyped for H1 and H3. At the peak of the 2022 Ottawa influenza testing, those samples represented only 18% of all tests performed. The remaining 82% of performed tests corresponded to samples collected from symptomatic hospitalized patients or symptomatic children admitted to the ER, which were sent to the Eastern Ontario Regional Laboratory Association (EROLA) where they are only typed for IAV or IBV. As a result, IAV subtyping via RT-qPCR WWS is near real-time and uses minimal resources and infrastructure creating unique opportunities for additional support of public health units/agencies to aid traditional clinical surveillance.

## Data Availability

All data produced will be available upon reasonable request to the authors, and will be shown on www.613covid.ca

## 4. Acknowledgements

The authors wish to acknowledge the help and assistance of the University of Ottawa, the Ottawa Hospital, the Children’ s Hospital of Eastern Ontario, the Children’ s Hospital of Eastern Ontario’ s Research Institute, Public Health Ontario, and all their employees involved in the project. The authors also wish to thank Dr. Earl Brown for providing the influenza virus stock. Their time, facilities, resources, and feedback are greatly appreciated.

