## Supplemental Files for "Wastewater surveillance of influenza activity: Early detection, surveillance, and subtyping in city and neighbourhood communities"

### 1. Site description and additional information

The four studied sites consisted of the City of Ottawa's Robert O. Pickard Environmental Centre, Ottawa's sole water resource recovery facility (WRRF), and three neighbourhoods, which were sampled by taking samples from manholes located in sewer collectors geographically isolating their wastewater. Additional information regarding population serviced, flows and some demographical information is shown below, in Table S1.

Table S1: Additional information for the 4 sites sampled during this study, including approximate population of the sewershed, number of included households in the sewershed, mean age of the population living within the sewershed, and average dry daily flow.

| | City of Ottawa WRRF | Neighbourhood #1 | Neighbourhood #2 | Neighbourhood #3 |
|---|---|---|---|---|
| Population (approx.) | 910,000 | 13,000 | 17,500 | 20,000 |
| # of households (approx.) | 396,000 | 5,200 | 3,200 | 2,300 |
| Mean age of population | 40.0 | 30.8 | 43.2 | 39.0 |
| Average daily flow ($m^3$/day) | 430,000 | 1,390 | 3,350 | 2,375 |

### 2. Probes and primers used in this study

All probes and primers used in this study, along with their manufacturer/place of origin, are shown below in Table S2.

Table S2: List of all probes and primers utilized in this study.

| Primer/probe (supplier) | Sequence | Reference |
|---|---|---|
| 2019-nCoV_N1 forward primer (IDT) | GAC CCC AAA ATC AGC GAA AT | [1] |
| 2019-nCoV_N1 reverse primer (IDT) | TCT GGT TAC TGC CAG TTG AAT CTG | [1] |
| 2019-nCoV_N1 probe (IDT) | 6-FAM-ACC CCG CAT /ZEN/ TAC GTT TGG TGG ACC-IBFQ | [1] |
| 2019-nCoV_N2 forward primer (IDT) | TTA CAA ACA TTG GCC GCA AA | [1] |
| 2019-nCoV_N2 reverse primer (IDT) | GCG CGA CAT TCC GAA GAA | [1] |
| 2019-nCoV_N2 probe (IDT) | 6-FAM-ACA ATT TGC /ZEN/ CCC CAG CGC TTC AG-IBFQ | [1] |
| PMMoV forward primer (ABI) | GAG TGG TTT GAC CTT AAC GTT GA | [2] |
| PMMoV reverse primer (ABI) | TTG TCG GTT GCA ATG CAA GT | [2] |
| PMMoV probe (ABI) | 6-FAM-CCT ACC GAA GCA AAT G-MGB | [2] |
| IAV M forward primer (ABI) | CAA GAC CAA TCY TGT CAC CTC TGA C | [1] |
| IAV M reverse primer (ABI) | GCA TTY TGG ACA AAV CGT CTA CG | [1] |
| IAV M probe (IDT) | 6-FAM-TGC AGT CCT /ZEN/ CGC TCA CTG GGC ACG-IBFQ | [1] |
| IBV NS forward primer (ABI) | TCC TCA AYT CAC TCT TCG AGC G | [1] |
| IBV NS reverse primer (ABI) | CGG TGC TCT TGA CCA AAT TGG | [1] |
| IBV NS probe (IDT) | 6-FAM-CCA ATT CGA /ZEN/ GCA GCT GAA ACT GCG GTG-IBFQ | [1] |
| H3N2 forward primer (ABI) | TGG ATT TCM TTY GCC ATA TCA T | [3] |
| H3N2 reverse primer (ABI) | GCA AAT GTT GCA YCT RAT RTT G | [3] |
| H3N2 probe (IDT) | 6-FAM-TGG CAR GCC CAC AT-MGB | [3] |
| H1N1 forward primer (ABI) | TGC TTT CGC ACT GAG TAG AGG | [4] |
| H1N1 reverse primer (ABI) | GGG CAC TCT CCT ATT GTG ACT G | [4] |

| | | |
|---|---|---|
| H1N1 probe (IDT) | 6-HEX-CCG GCA TCA TCA CCT CAA ACG CRT CAA-BHQ1 | [4] |
| Stuffer-1 forward primer | CAA AGC GAG AAC GGA TCC GGA GCC ACG AAC TTC CAA GCA GGA GAC | This study |
| Stuffer-2 forward primer | ACC TGC CAA CCA AAG CGA GAA C | This study |
| IAV seq-Stuffer forward primer | CAA GCA GGA GAC GTG GAA GAA AAC CCC GGT CCT CAA GAC CAA TC | This study |

#### 3. RT-qPCR analysis and cycling conditions

All PCR reactions performed in this study were singleplex, TaqMan one-step RT-qPCR experiments. Samples were run in triplicate using a CFX96 touch real time thermocycler (Bio-Rad, Hercules, CA). PCR cycling conditions for all assays performed in this study are described below in detail in Table S3.

The limit of detection of the RT-qPCR assay for influenza M was determined by evaluating the least concentration(copies) per reaction being detected with a detection rate of ≥95% (<5% false negatives), as recommended by the MIQE guidelines[5]. Furthermore, samples were discounted if the following experimental conditions were not met: i) the standard curves have an $R^2 \geq 0.95$, ii) the copies/reaction are in linear dynamic range of the standard curve and iii) the primer efficiency lies between 90% to 120%. Furthermore, sample replicates with values greater than 0.5 standard deviations away from the average of the triplicates discounted.

Table S3. List of RT-qPCR conditions for all targets investigated in this study, including thermal cycling conditions, primer and probe concentrations, and QA/QC controls employed during this study.

| | Targets | | |
|---|---|---|---|
| | N1, N2, PMMoV | IAV, IBV | H3N2, H1N1 subtyping |
| **RT-qPCR conditions** | Reverse trans.: 5 min. @ 50°C, 1 cycle<br>Initial denat.: 20 sec. @ 95°C, 1 cycle<br>Denaturation: 3 sec. @ 95°C, 45 cycles<br>Anneal/ext.: 30 sec. @ 60°C, 45 cycles | Reverse trans.: 5 min. @ 50°C, 1 cycle<br>Initial denat.: 20 sec. @ 95°C, 1 cycle<br>Denaturation: 3 sec. @ 95°C, 45 cycles<br>Anneal/ext.: 30 sec. @ 60°C, 45 cycles | Reverse trans.: 5 min. @ 50°C, 1 cycle<br>Initial denat.: 20 sec. @ 95°C, 1 cycle<br>Denaturation: 3 sec. @ 95°C, 45 cycles<br>Anneal/ext.: 30 sec. @ 56.3/57°C, 45 cycles |
| **Primer and probe concentrations** | 500 µM (primers)<br>125 µM (probes) | 500 µM (primers)<br>200 µM (probes) | 500 µM (primers)<br>200 µM (probes) |
| **Supermix used and total reaction volume** | 1-Step Fast Virus (2.5 µL), 10 µL | 1-Step Fast Virus (2.5 µL), 10 µL | 1-Step Fast Virus (2.5 µL), 10 µL |
| **Replicate exclusion threshold** | Ct ≥ 0.5 | Ct ≥ 0.5 | Ct ≥ 0.5 |
| **Standard curve acceptance characteristics** | $R^2 \geq 0.95$,<br>90% ≤ Eff. ≤ 120% | $R^2 \geq 0.95$,<br>90% ≤ Eff. ≤ 120% | $R^2 \geq 0.95$,<br>90% ≤ Eff. ≤ 120% |
| **Other QA/QC performed** | Extraction blank, no extrapolation of values, negative controls | Extraction blank, no extrapolation of values, negative controls | Extraction blank, no extrapolation of values, negative controls |

#### 4. RT-ddPCR analysis and cycling conditions

Singleplex, probe-based, one-step RT-ddPCR was used for absolute quantification of the material used as standards for the assays used in this study. All primers and probes used in this assay are shown above in Supplemental Table 2. Analysis was performed with a reaction volume of 20 µL, which consisted of 5 µL of RNA template, 5 µL of 4x One-Step RT-ddPCR Advanced Kit for Probes (Bio-Rad, CA, USA), 900 nM of both forward and reverse primers and 250 nM of the probe, with nuclease-free water making up the balance. Samples were prepared and analyzed in triplicate. The reaction was emulsified using a QX200 droplet generator (Bio-Rad, CA, USA) and droplets were transferred to a new ddPCR reaction plate. PCR was completed using a C1000 Touch™ Thermal Cycler with 96 deep wells (Bio-Rad, CA, USA) under the following conditions: reverse transcriptase (RT) was performed at 50°C (60 minutes), followed by polymerase activation at 95°C (10 minutes), followed by 40 cycles of denaturation at 94°C (30 seconds) and annealing/extension 55°C (60 seconds), respectively. The polymerase was finally deactivated by holding 98°C for 10 minutes, followed by a droplet stabilization period at 4°C for 30 minutes. Droplets were then read using a QX200 droplet reader (Bio-Rad, CA, USA). Positive droplets were called manually, and absolute quantification was performed using the Quantasoft Analysis Pro software (ver. 1.0) (Bio-Rad, CA, USA).

#### 5. Sanger sequencing of amplicons

The specificity of amplicons generated for various targets in this study was evaluated via Sanger sequencing of the DNA amplicons resulting from RT-qPCR analysis. First, a touchdown PCR (TD-PCR) was performed using Q5® High-Fidelity DNA Polymerase with 1 µl of RT-qPCR amplicons as the starting template. The initial touchdown was performed as follows: [98°C (30 seconds) + 64°C → 55˚C, drop of 1°C/cycle, + 72°C (30 seconds)] x 10 cycles. Amplification was then performed as follows: [98°C (30 seconds) + 64°C → 55˚C, drop of 0.4°C/cycle, + 72°C (30 seconds)] x 25 cycles. The TD-PCR products were then run on a 3% agarose gel at 100V to separate the amplicons. The amplicon band observed at the appropriate location (the IAV M amplicon was located at 106 base pair) was then cut and gel extracted using Monarch® DNA Gel Extraction Kit (New England Biolabs, MA, USA) as per the manufacturer's instructions. After obtaining purified DNA, a novel primer extension strategy for one-step PCR amplification was performed using the Q5® High-Fidelity DNA Polymerase. In brief, 1 ng of DNA was used to extend the IAV M amplicon using the oligo adapters (Supplemental Table 1) with partial complementarity to the targeted amplicons. An extension PCR amplification was then performed as follows: 98°C (30 seconds) + [98°C (10 seconds) + 50°C (30 seconds) + 72°C (30 seconds) x 30 cycles, + 72°C (2 minutes). The extension-PCR amplified product is then purified using QIAquick® PCR Purification Kit (Qiagen, MD, USA) following the manufacturer's instructions. The final amplicon product was then sequenced by Sanger Sequencing at the Ottawa Hospital's Research Institute (OHRI) StemCore Sequencing Facility using an ABI Prism 3730 DNA Sequencer (Applied Biosystems, MA, USA). The sequences were compiled and edited using BioEdit (ver. 7.2)[6] and sequences alignment was done by Clustal Omega[7]. Each PCR reaction had a total volume of 25 µl and was composed of the amplicons' regular reverse primers (500 nM), A target-specific amplicon-seq-Stuffer forward primer (50 nM), Stuffer-1 forward primer (50 nM), Stuffer-2 forward primer (500 nM), dNTP (200 µM), and 1X of the 5X Q5 reaction buffer and Q5 high fidelity DNA polymerase (0.02U/µl).

#### 6. Standard material preparation for IAV and IBV

To assess the reaction efficiency and limit of detection (LOD) of our RT-qPCR assay, cell cultured HK/1/68/MA/E2 (H3N2) mouse adapted virus strain and WSN/33 (H1N1) virus strain were used to extract RNA. Viral RNA was ddPCRed and subject to a serial dilution. One-step RT-qPCR was performed for those viral RNA with validated primers/probe set. LOD was calculated based on the LOD definition.

RT-ddPCR quantified the influenza A viral RNA was used as standard for quantifying wastewater RNA samples. A serial dilution of viral RNA standard was included on every 96-well PCR plate to produce standard curves used to quantify the copies of influenza viral genes. Additionally, RT-qPCR runs were validated with the use of non-template-controls (NTCs), positive controls, negative controls of pre-influenza season wastewater samples and dilutions.
